# Epidemiological, Clinical Characteristics and Outcome of Medical Staff Infected with COVID-19 in Wuhan, China: A Retrospective Case Series Analysis

**DOI:** 10.1101/2020.03.09.20033118

**Authors:** Jie Liu, Liu Ouyang, Pi Guo, Haisheng Wu, Peng Fu, Yuliang Chen, Dan Yang, Xiaoyu Han, Yukun Cao, Osamah Alwalid, Juan Tao, Shuyi Peng, Heshui Shi, Fan Yang, Chuansheng Zheng

**Affiliations:** Department of Radiology, Union Hospital, Tongji Medical College, Huazhong University of Science and Technology, Wuhan, Hubei, China; Hubei Province Key Laboratory of Molecular Imaging, Wuhan, Hubei, China; Department of Orthopaedics, Union Hospital, Tongji Medical College, Huazhong University of Science and Technology, Wuhan, Hubei, China; Department of Preventive Medicine, Shantou University Medical College, Shantou, Guangdong, China; Department of Thyroid and Breast Surgery, The Central Hospital of Wuhan, Tongji Medical College, Huazhong University of Science and Technology, Wuhan, Hubei, China; Department of Respiratory and Critical Care Medicine, Union Hospital, Tongji Medical College, Huazhong University of Science and Technology, Wuhan, Hubei, China

**Keywords:** COVID-19, medical staff, infectious disease, retrospective study, Prognosis

## Abstract

**Backgrounds:** Since December 2019, a novel coronavirus epidemic has emerged in Wuhan city, China and then rapidly spread to other areas. As of 20 Feb 2020, a total of 2,055 medical staff confirmed with coronavirus disease 2019 (COVID-19) caused by SARS-Cov-2 in China had been reported. We sought to explore the epidemiological, clinical characteristics and prognosis of novel coronavirus-infected medical staff.

**Methods:** In this retrospective study, 64 confirmed cases of novel coronavirus-infected medical staff admitted to Union Hospital, Wuhan between 16 Jan, 2020 to 15 Feb, 2020 were included. Two groups concerned were extracted from the subjects based on duration of symptoms: group 1 (≤10 days) and group 2 (>10 days). Epidemiological and clinical data were analyzed and compared across groups. The Kaplan-Meier plot was used to inspect the change in hospital discharge rate. The Cox regression model was utilized to identify factors associated with hospital discharge.

**Findings:** The median age of medical staff included was 35 years old. 64% were female and 67% were nurses. None had an exposure to Huanan seafood wholesale market or wildlife. A small proportion of the cohort had contact with specimens (5%) as well as patients in fever clinics (8%) and isolation wards (5%). Fever (67%) was the most common symptom, followed by cough (47%) and fatigue (34%). The median time interval between symptoms onset and admission was 8.5 days. On admission, 80% of medical staff showed abnormal IL-6 levels and 34% had lymphocytopenia. Chest CT mainly manifested as bilateral (61%), septal/subpleural (80%) and ground-glass (52%) opacities. During the study period, no patients was transferred to intensive care unit or died, and 34 (53%) had been discharged. Higher body mass index (BMI) (≥ 24 kg/m^2^) (HR 0.14; 95% CI 0.03-0.73), fever (HR 0.24; 95% CI 0.09-0.60) and higher levels of IL-6 on admission (HR 0.31; 95% CI 0.11-0.87) were unfavorable factors for discharge.

**Interpretation:** In this study, medical staff infected with COVID-19 have relatively milder symptoms and favorable clinical course, which may be partly due to their medical expertise, younger age and less underlying diseases. Smaller BMI, absence of fever symptoms and normal IL-6 levels on admission are favorable for discharge for medical staff. Further studies should be devoted to identifying the exact patterns of SARS-CoV-2 infection among medical staff.

## Introduction

In December, 2019, a group of novel atypical pneumonia patients with uncertain etiology but mostly linked to the Huanan Seafood Wholesale Market emerged in Wuhan, China [1]. A later confirmed pathogen of this previously unknown pneumonia was described as a novel coronavirus, currently named as severe acute respiratory syndrome coronavirus 2 (SARS-Cov-2; previously known as 2019nCoV), was ascertained by unbiased sequencing analysis of lower respiratory tract samples from early cases on 7 Jan 2020, following which the protocol of real-time reverse-transcriptase polymerase chain reaction (RT-PCR) assay for this novel coronavirus had also been developed [2-7]. In fact, the epidemics of the two other novel coronaviruses, namely severe acute respiratory syndrome coronavirus (SARS-CoV) and Middle East respiratory syndrome coronavirus (MERS-CoV), have posed a huge threat to public health in the past two decades [8, 9]. SARS-Cov-2 in this outbreak, like the previous two viruses, is also categorized within the same genus of the subfamily Orthocoronavirinae within the family Coronaviridae, but shows a strong affinity for human respiratory receptors [10, 11].

By 11 Feb 2020, coronavirus disease 2019 (COVID-19) due to the SARS-Cov-2 has caused more than 40,000 laboratory confirmed cases and 1,023 deaths among them in China [12]. Sufficient evidence indicated that the COVID-19 clustered within close-contact human groups, such as family and hospital settings [13-17]. The SARS-CoV-2 epidemic has transmitted throughout China and to other countries due to massive population movements before the Lunar New Year [14], and consequently escalated as a Public Health Emergency of International Concern declared by World Health Organization (WHO) [18]. As of 4 Mar 2020, more than 90,000 confirmed cases infected with SARS-CoV-2 have been identified globally [19].

Information pointing to the epidemiology and clinical features of general confirmed cases has been accumulating. The previous studies enrolling 41, 99 and 138 confirmed cases admitted to Wuhan, respectively, provided an insight into epidemiological characteristics, clinical manifestations, treatment measures and clinical outcomes of these patients [1, 20, 21]. In particular, a recent study in Zhejiang province, China indicated that the symptoms of patients outside of Wuhan perhaps are relatively mild versus symptoms of initial cases in Wuhan [10]. Meanwhile, a new finding from a national wide descriptive report drew a huge amount of attention, which declared that the total number of confirmed novel coronavirus-infected medical staff was as high 1,716 as of 11 February 2020, with a peak incidence occurring on 28 January 2020 [12]. Hospital-related transmission are one of the causes for infection of health-care workers [21], especially in the early stages of COVID-19 epidemic when there was a lack of knowledge about transmission approaches of SARS-CoV-2, as well as in the period when facing a shortage of protective materials. Nonetheless, the predominant cause of the infection and the failure of protection among health workers remains to be investigated [12].

Despite the increased attention towards protecting medical staff from infection, information regarding the epidemiology and clinical features of medical staff confirmed with COVID-19 is scarce. This single-centered, retrospective study aimed to describe epidemiological, clinical, laboratory and radiographic features, treatment, and prognosis of a group of medical staff confirmed with COVID-19 who were admitted to Union Hospital, Wuhan. We hope the findings in the present study will provide an insight into the prevention and treatment of this novel coronavirus for the global community.

## Methods

### Study design and participants

We performed a single-centered, retrospective study on a group of novel coronavirus-infected medical staff at Wuhan Union Hospital, one of the hospitals treating patients confirmed with COVID-19 at the earliest time. Diagnosis of cases with SARS-Cov-2 infection conforms to the WHO interim guidance [7]. Details regarding laboratory confirmation protocol for SARS-CoV-2 were described by previous studies [1, 21]. Throat-swab specimens were screened for SARS-CoV-2 and other respiratory viruses (influenza, respiratory syncytial virus, etc.) by real-time RT-PCR assays. A total 64 medical staff, who were confirmed by SARS-CoV-2 real-time RT-PCR test on respiratory secretions collected by throat swab and undergone serial chest CT scans following their admission to isolation wards of Union Hospital between 16 Jan and 15 Feb, 2020, were enrolled.

This retrospective study was approved by the Ethics of Committees of Union Hospital, Tongji Medical College, Huazhong University of Science and Technology. Written informed consent was waived due to the rapid emergence of this infectious disease.

### Data collection

The epidemiological data, medical and nursing records, laboratory examinations, chest computed tomography (CT) of all patients were reviewed and abstracted with concerted efforts of experienced clinicians. Data were collected at the time of symptoms onset, presentation for medical advice and in-patient admission. The clinicians who had experience of treating patients with confirmed SARS-Cov-2 infection reviewed and collected the medical records of patients, and preliminarily collated the data. The clinical data were extracted through a standardized form for case report as previously described [23]. Epidemiological data, including exposure histories before symptoms onset (whether there is a history of exposure to the Huanan Seafood Wholesale Market, or wildlife), and close contact with laboratory-confirmed or suspected cases of COVID-19 in a work environment (fever clinics, or isolation wards), specimens (pharyngeal swab, blood, sputum specimens, etc.) or family members with COVID-19 were collected. In addition, information about preventive medication among medical staff was also collected.

We have also collected the data on demographics, clinical manifestations, laboratory examinations and radiological studies. These included age, sex, occupation (doctor, or nurse), body mass index (BMI ≥24, or <24 kg/m^2^), current smoking status (yes, or no), disease severity (non-severe, or severe), date of symptom onset, symptoms before hospital admission (fever, cough, fatigue, sore throat, myalgia, sputum production, difficulty breathing or chest tightness, chill, loss of appetite, diarrhea, and chest pain), coexisting conditions (e.g. hypertension, diabetes, etc.), laboratory testing indicators on admission (leucocyte count, lymphocyte count, platelet count, D-dimer, creatinine, creatine kinase, lactose dehydrogenase, alanine aminotransferase, aspartate aminotransferase, hemoglobin, ferritin, C-reactive protein, Amyloid A, total bilirubin, procalcitonin, erythrocyte sedimentation rate, interleukin-6 (IL-6) and lymphocyte subsets, etc.), radiologic assessments of chest CT (lung involvement, lung lobe involvement, predominant CT changes, predominant distribution of opacities, etc.), treatment measures (antibiotics agents, antiviral agents, traditional Chinese medicine, immune globulin, thymosin, corticosteroids and oxygen therapy), and complications (e.g. pneumonia, acute respiratory distress syndrome, acute cardiac injury, acute kidney injury, shock, etc.). All CT images were analyzed by two radiologists (J.L. and F.Y., who had 5 and 21 years of experience in thoracic radiology, respectively) utilizing the institutional digital database system without access to clinical and laboratory findings. Images were reviewed independently, and final decisions were reached by discussion and consensus. We estimated the time interval from symptom onset to admission with maximum information available - that is, all the exact date of initial symptoms provided by the patients. Then the aggregated data was sent to data analysis group. Prior to statistical analysis, the aggregated data were cross - checked by group members to guarantee the correctness and completeness of data.

### Outcomes

The clinical outcomes and prognosis were continuously observed up to 24 Feb 2020. We defined the primary outcomes as discharge. The discharge criteria of inpatients included all the following three conditions [24]: (1) body temperature return to normal for more than 3 days and respiratory symptoms improvement; (2) resolution of lung involvement demonstrated by chest CT; (3) two consecutive RT-RCR tests, with sampling interval of more than 1 day, showing a negative result. Secondary outcomes consisted of hospital discharge rate and length of hospital stay. Given that treatment and monitoring of some patients in our study were still ongoing, a fixed time-interval of observation was not applied to these clinical outcomes.

### Statistical analysis

This study devoted to report the epidemiological, clinical characteristics and prognosis of medical staff confirmed with COVID-19. Continuous variables were checked for distribution normality by means of the Kolmogorov-Smirnov test, following which they were summarized as either means with standard deviations (SD) or medians with interquartile ranges (IQR) as appropriate. Counts and percentages were utilized to describe categorical variables. Given the cut-off point at 10th day of symptoms onset proposed by previous studies [1, 21], we assigned the patients into either one of two groups based on duration of symptoms: group 1 (≤10 days) and group 2 (>10 days). We applied a Kaplan-Meier plot to inspect the change of hospital discharge rate. The proportional hazard Cox regression model was utilized to ascertain factors associated with hospital discharge. Univariate models with a single variable once at a time were first fitted. The statistically significant risk factors as well as age and sex were, then, included into a final multivariate Cox regression model. The hazards ratios (HRs) along with the 95% confidence intervals (95% CIs) were calculated.

Statistical tests were two-sided with significance set at α less than 0.05. We performed all data analyses by R software version 3.6.2 (R Foundation for Statistical Computing).

## Results

### Epidemiological characteristics

During the study period, epidemiological and clinical data were collected on 64 medical staff with laboratory-confirmed SARS-Cov-2 infection from Wuhan Union Hospital, of whom 62 (97%) provided an exact date of symptom onset and only 1 case (2%) was severe. The patients aged between 23 and 63 years old, and median age was 35 years (IQR 29-43 years). The median age in group 1 was 37 years (IQR 32-44 years), and in group 2 it was 30 years (IQR 27-36 years). More than half of the cohort were female (64%) and nurse (67%). There were 7 (11%) overweight cases (BMI ≥ 24 kg/m^2^) and only 1 was current smoker.

Among the 64 medical staff recruited, no one had an exposure to Huanan seafood wholesale market or wildlife, while 4 (6%) medical staff had family members confirmed with SARS-CoV-2 infection. During patient care, 5 (8%) and 3 (5%) cases had contact with patients in fever clinics and isolation wards, respectively, and 3 (5%) had direct contact with specimens collected from confirmed patients. 10 (16%) of 64 medical staff have used preventive medications (Table 1).

**Table 1:**
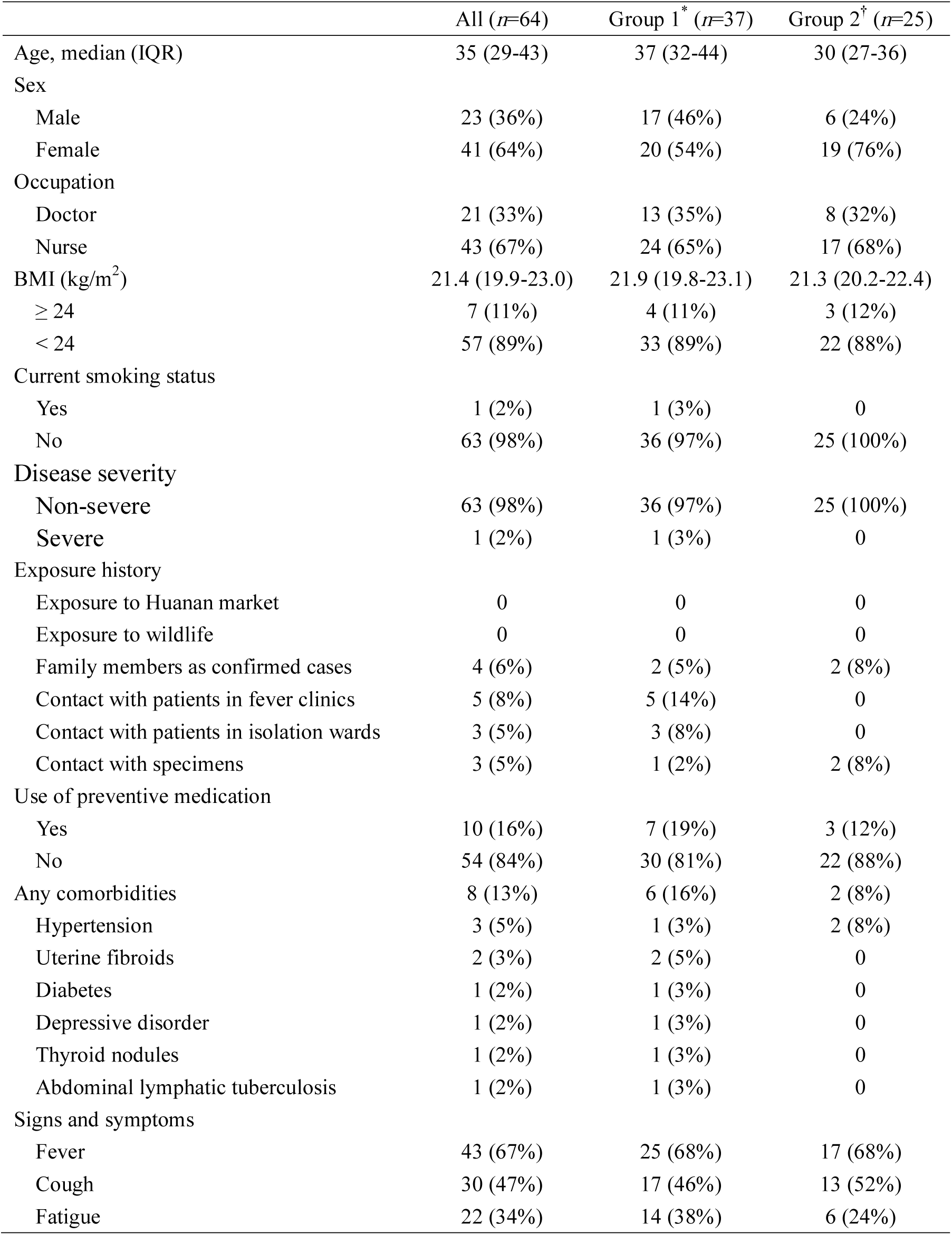
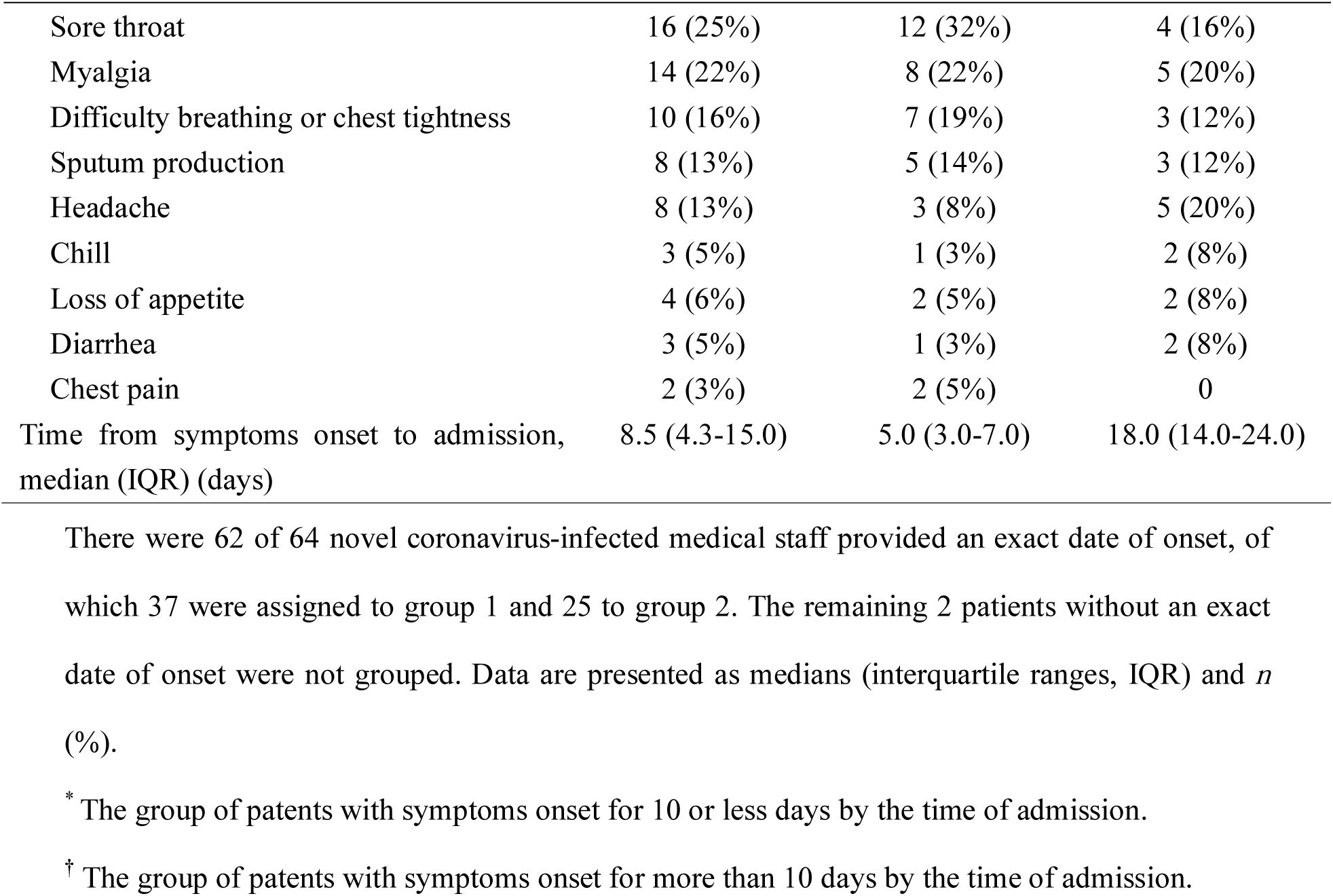
Demographics and baseline characteristics of 64 medical staff infected with COVID-19 pneumonia in Wuhan, China.

### Clinical features

The median duration between symptoms onset and admission was 8.5 (IQR 4.3-15.0) days of the entire cohort; 5.0 (IQR 3.0-7.0) days in group 1 and 18.0 (IQR 14.0-24.0) days in group 2. There were 8 (13%) cases, most of whom were assigned to group 1, with one or more co-morbidities: 3 (5%) had hypertension, 2 (3%) had uterine fibroids, and one (2%) each had diabetes, depressive disorder, thyroid nodules or abdominal lymphatic tuberculosis. The three most common symptoms were fever (67%), cough (47%) and fatigue (34%). The relatively less common symptoms were sore throat, myalgia, difficulty breathing or chest tightness, sputum production, headache, chill, loss of appetite, diarrhea, and chest pain (Table 1).

Table 2 shows the laboratory and radiographic findings of 64 medical staff with confirmed COVID-19. On admission, the blood counts of 11 (17%) cases showed leukocytopenia and only one (2%) showed leukocytosis. 22 (34%) presented with lymphocytopenia and 7 (11%) presented thrombocytopenia. Most cases demonstrated normal levels of D-dimer, creatinine, and creatine kinase, but elevated C-reactive protein and amyloid A levels were presented in 45% and 59% of cases, respectively. Elevated levels of alanine aminotransferase (13%) and aspartate aminotransferase (9%) were less common. A small proportion (3%) of cases had abnormal procalcitonin serum level (>0.5 ug/L). Notably, 47 (80%) of cases had high levels of IL-6 (>2.9 pg/ml). Medical staff of group 1 had more prominent laboratory abnormalities (i.e., leukocytes, lymphocytes, platelet, alanine aminotransferase, amyloid A and IL-6) as compared with those in group 2.

**Table 2:** Laboratory and radiographic findings of 64 medical staff infected with COVID-19 pneumonia in Wuhan, China.

|  | Normal range | All (n=64) | Group 1* (n=37) | Group 2† (n=25) |
| --- | --- | --- | --- | --- |
| Leukocytes ($\times 10^9$/L) | 3.5-9.5 | 4.7 (3.8-5.7) | 4.0 (3.4-5.0) | 5.7 (4.7-6.6) |
| Decreased |  | 11 (17%) | 10 (27%) | 1 (4%) |
| Increased |  | 1 (2%) | 0 | 1 (4%) |
| Neutrophilic granulocyte percentage (%) | 40-75 | 56.9 (51.4-66.3) | 56.3 (50.3-66.0) | 57.5 (51.5-65.1) |
| Lymphocytes ($\times 10^9$/L) | 1.1-3.2 | 1.4 (0.9-1.8) | 1.3 (0.9-1.6) | 1.6 (1.1-2.2) |
| Decreased |  | 22 (34%) | 14 (38%) | 7 (28%) |
| Platelets ($\times 10^9$/L) | 115-350 | 188 (148-211) | 160 (130-201) | 195 (178-247) |
| Decreased |  | 7 (11%) | 6 (16%) | 1 (4%) |
| D-dimer (mg/L) | 0-0.5 | 0.2 (0.2, 0.4) | 0.2 (0.2, 0.4) | 0.2 (0.2, 0.4) |
| Creatinine (umol/L) | 44-106 | 66.6 (59.1-80.5) | 68.3 (59.9-86.9) | 65.8 (59.1-73.1) |
| Creatine kinase (U/L) | 26-140 | 60.0 (45.5-91.5) | 62.0 (47.5-104) | 60.0 (44.0-77.0) |
| Lactose dehydrogenase (U/L) | 109-245 | 189 (170-209) | 190 (177-208) | 185 (154-210) |
| Alanine aminotransferase (U/L) | 5-35 | 17.5 (13.8-29.5) | 20.0 (15.0-31.0) | 15.0 (13.0-27.0) |
| Increased |  | 8 (13%) | 6 (16%) | 2 (8%) |
| Aspartate aminotransferase (U/L) | 8-40 | 21.5 (16.0-27.3) | 23.0 (17.0-29.0) | 17.0 (15.0-23.0) |
| Increased |  | 6 (9%) | 5 (14%) | 1 (4%) |
| Hemoglobin (g/L) | 115-150 | 130 (119-141) | 133 (124-142) | 125 (119-136) |
| Ferritin (ug/L) | 4.6-204 | 103 (62-297) | 145 (79-448) | 90 (50-133) |
| C-reactive protein >8 mg/L | 0-8 | 29 (45%) | 19 (51%) | 9 (36%) |
| Amyloid A (mg/L) | 0-10 | 25.7 (4.0-150.0) | 72.4 (16.9-176) | 3.8 (2.3-35.9) |
| Increased |  | 34/58 (59%) | 25/32 (78%) | 7/24 (29%) |
| Procalcitonin >0.5 ug/L | 0-0.5 | 2/61 (3%) | 2/35 (6%) | 0 |
| Total bilirubin (umol/L) | 5.1-19 | 10.0 (8.2-13.5) | 9.5 (7.9-12.5) | 12.0 (8.2-14.9) |
| Urea nitrogen (mmol/L) | 2.9-8.2 | 3.9 (3.0-4.9) | 3.9 (3.0-5.1) | 4.0 (3.5-4.7) |
| Erythrocyte sedimentation rate (mm/h) | 0-20 | 10.0 (6.0-24.0) | 13.0 (6.0-25.0) | 7.5 (4.8-16.0) |
| IL-6 (pg/ml) | 0.1-2.9 | 4.4 (3.4-6.6) | 4.7 (4.1-7.4) | 3.3 (2.7-4.3) |
| Increased |  | 47/59 (80%) | 32/35 (91%) | 13/22 (59%) |
| lymphocyte subsets |  |  |  |  |
| CD3+ ratio (%) | 58.17-84.22 | 76.5 (70.5-80.4) | 73.2 (70.0-79.6) | 79.0 (74.4-82.0) |
| CD4+ ratio (%) | 25.34-51.37 | 42.9 (37.9-48.5) | 41.6 (37.6-43.6) | 45.9 (38.1-51.5) |
| CD8+ ratio (%) | 14.23-38.95 | 26.8 (23.6-30.2) | 26.3 (22.8-30.7) | 26.8 (24.8-29.6) |
| B-CELL ratio (%) | 4.1-18.31 | 11.0 (8.9-14.8) | 10.1 (8.3-14.7) | 12.1 (10.0-15.3) |
| NK cell ratio (%) | 3.33-30.47 | 6.2 (4.1-10.8) | 8.6 (5.4-11.9) | 4.6 (2.5-6.3) |
| Ratio of CD4/CD8 | 0.41-2.72 | 1.6 (1.3-2.0) | 1.6 (1.3-2.0) | 1.5 (1.3-2.1) |
| Abnormalities on chest CT | - | 58 (91%) | 35 (95%) | 21 (84%) |
| Lung involvement |  |  |  |  |
| Unilateral | - | 19 (30%) | 11 (30%) | 7 (28%) |
| Bilateral | - | 39 (61%) | 24 (65%) | 14 (56%) |
| Lung lobe involved |  |  |  |  |
| Right upper lobe | - | 20 (31%) | 14 (38%) | 5 (20%) |
| Right middle lobe | - | 16 (25%) | 11 (30%) | 5 (20%) |
| Right lower lobe | - | 45 (70%) | 27 (73%) | 17 (68%) |
| Left upper lobe | - | 22 (34%) | 15 (41%) | 7 (28%) |
| Left lower lobe | - | 41 (64%) | 27 (73%) | 13 (52%) |
| Predominant CT pattern |  |  |  |  |
| Ground glass opacity | - | 33 (52%) | 27 (46%) | 14 (56%) |
| Consolidation | - | 8 (13%) | 6 (16%) | 2 (8%) |
| Mixed pattern | - | 17 (27%) | 12 (32%) | 5 (20%) |
| Predominant distribution of opacities |  |  |  |  |
| Septal/subpleural | - | 51 (80%) | 30 (81%) | 19 (76%) |
| Peribronchovascular | - | 3 (5%) | 2 (5%) | 1 (4%) |
| Random | - | 4 (6%) | 3 (8%) | 1 (4%) |
| Thickening of the adjacent pleura | - | 3 (5%) | 2 (5%) | 1 (4%) |
| Pleural effusion |  | 0 | 0 | 0 |
| Lymphadenopathy | - | 0 | 0 | 0 |
| Any original chronic lung lesion | - | 19 (30%) | 10 (27%) | 8 (32%) |

There were 62 of 64 novel coronavirus-infected medical staff provided an exact date of onset, of which 37 were assigned to group 1 and 25 to group 2. The remaining 2 patients without an exact date of onset were not grouped. Data are presented as medians (interquartile ranges, IQR) and *n* (%). For each item, the effective sample size of total population, group 1 and group 2 is 64, 37 and 25 unless stated otherwise.
\* The group of patents with symptoms onset for 10 or less days by the time of admission.
† The group of patents with symptoms onset for more than 10 days by the time of admission.

As evidenced by Table 2 which illustrate the radiological findings in the study cohort on chest CT, 58 (91%) of 64 cases showed abnormalities (Figure 1). 39 (61%) had bilateral lung involvement (Figure 1, A and C). The right lower lobe (70 %) and left lower lobe (64%) were the most common involved lobes. Ground glass opacity was regarded as the predominant abnormality on chest CT (Figure 1, A and B) observed in 33 (52%) of cases, meanwhile subpleural distribution was predominant as identified in 51 (80%) (Figure 1, C). Thickening of the adjacent pleura, nodules, emphysema, pleural effusion and lymphadenopathy were relatively rare. CT scans also found that 19 (30%) of medical staff had one or more chronic lung lesions with a static picture on serial CT examinations (Figure 1, D).

**Figure 1:**
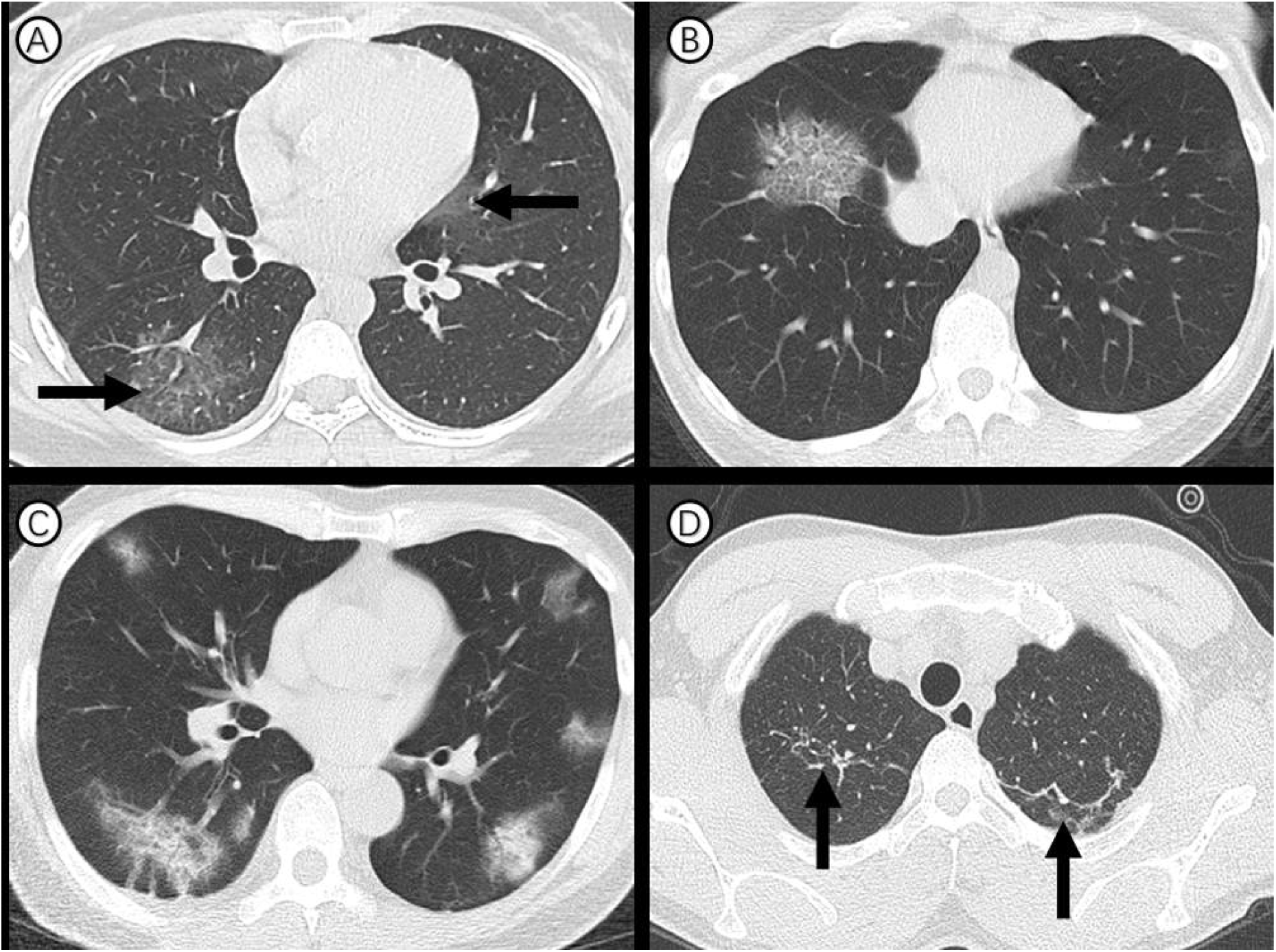
Transverse thin-section CT scans in medical staffs infected with COVID-19 pneumonia. (A) 27-year-old woman: bilateral, peripheral ground-glass opacity in the right lower lobe and left upper lobe (arrow). (B) 41-year-old woman: focal ground-glass opacity associated with smooth interlobular and intralobular septal thickening (crazy-paving pattern). (C) 55-year-old man: bilateral, peripheral ground-glass opacity mixed consolidation pattern. (D) 28-year-old man: bilateral and linear consolidations in the right and left upper lobes (arrow), regarded as chronic lung lesion with lack of changes on serial CT examinations.

### Treatment measures and prognosis

Of the study participants, no person was transferred to an intensive care unit for mechanical ventilation due to acute respiratory distress syndrome. 9 (14%) patients needed an electrocardiograph monitoring, among whom 8 were in group 1. Empirical intravenous antibiotic treatment was administered in 55 (86%) patients. All the patients were given empirical antiviral therapy. Meanwhile, 13 (20%) were offered traditional Chinese medicine, 23 (36%) patients were given immune globulin, 33 (52%) were given thymosin, and 7 (11%) received corticosteroids. As for oxygen therapy, 32 (50%) used nasal cannula and only 2 (3%) used face mask, while no one needed invasive mechanical or ventilation extracorporeal membrane oxygenation. As a whole, despite the negligible difference of antiviral treatment between two groups, most of the cases who received electrocardiogram monitoring, antibiotics, immune globulin, thymosin and oxygen therapy belonged to group 1, whereas the proportion given traditional Chinese medicine was higher in group 2 (Table 3).

**Table 3:** Treatments and outcomes of 64 medical staff infected with COVID-19 pneumonia in Wuhan, China.

|  | All (n=64) | Group 1<sup>*</sup> (n=37) | Group 2<sup>†</sup> (n=25) |
| --- | --- | --- | --- |
| Electrocardiograph monitoring | 9 (14%) | 8 (22%) | 1 (4%) |
| Antibiotics treatment | 55 (86%) | 37 (100%) | 17 (68%) |
| Antiviral treatment | 64 (100%) | 37 (100%) | 25 (100%) |
| Traditional Chinese medicine | 13 (20%) | 5 (14%) | 7 (28%) |
| Immune globulin | 23 (36%) | 18 (49%) | 5 (20%) |
| Thymosin | 33 (52%) | 20 (54%) | 12 (48%) |
| Corticosteroids | 7 (11%) | 5 (14%) | 2 (8%) |
| Oxygen therapy |  |  |  |
| Nasal cannula | 32 (50%) | 22 (59%) | 10 (40%) |
| Face mask | 2 (3%) | 2 (5%) | 0 |
| Length of hospital stay (days) | 12.5 (9.0-19.8) | 18.0 (13.0-20.5) | 9.0 (6.0-11.0) |
| Outcome |  |  |  |
| Hospital discharge | 34 (53%) | 19 (51%) | 13 (52%) |
| Continued hospitalization | 30 (47%) | 18 (49%) | 12 (48%) |
| Death | 0 | 0 | 0 |

There were 62 of 64 novel coronavirus-infected medical staff provided an exact date of onset, of which 37 were assigned to group 1 and 25 to group 2. The remaining 2 patients without an exact date of onset were not grouped. Data are presented as medians (interquartile ranges, IQR) and *n* (%).
<sup>\*</sup> The group of patients with symptoms onset for 10 or less days by the time of first admission.
<sup>†</sup> The group of patients with symptoms onset for more than 10 days by the time of first admission.

By 24 Feb, 2020, 34 (53%) of the cases have been discharged and none had died, the remaining cases were still in hospital to receive supportive therapy. The median length of hospital stay was 12.5 (IQR 9.0-19.8) days in total, 18.0 (IQR 13.0-20.5) days in group 1 and 9.0 (IQR 6.0-11.0) days in group 2 (Table 3). The overall median discharge time (i.e. equal to the time that half of the patients left the hospital) was 20 days (Figure 2A).

**Figure 2.**
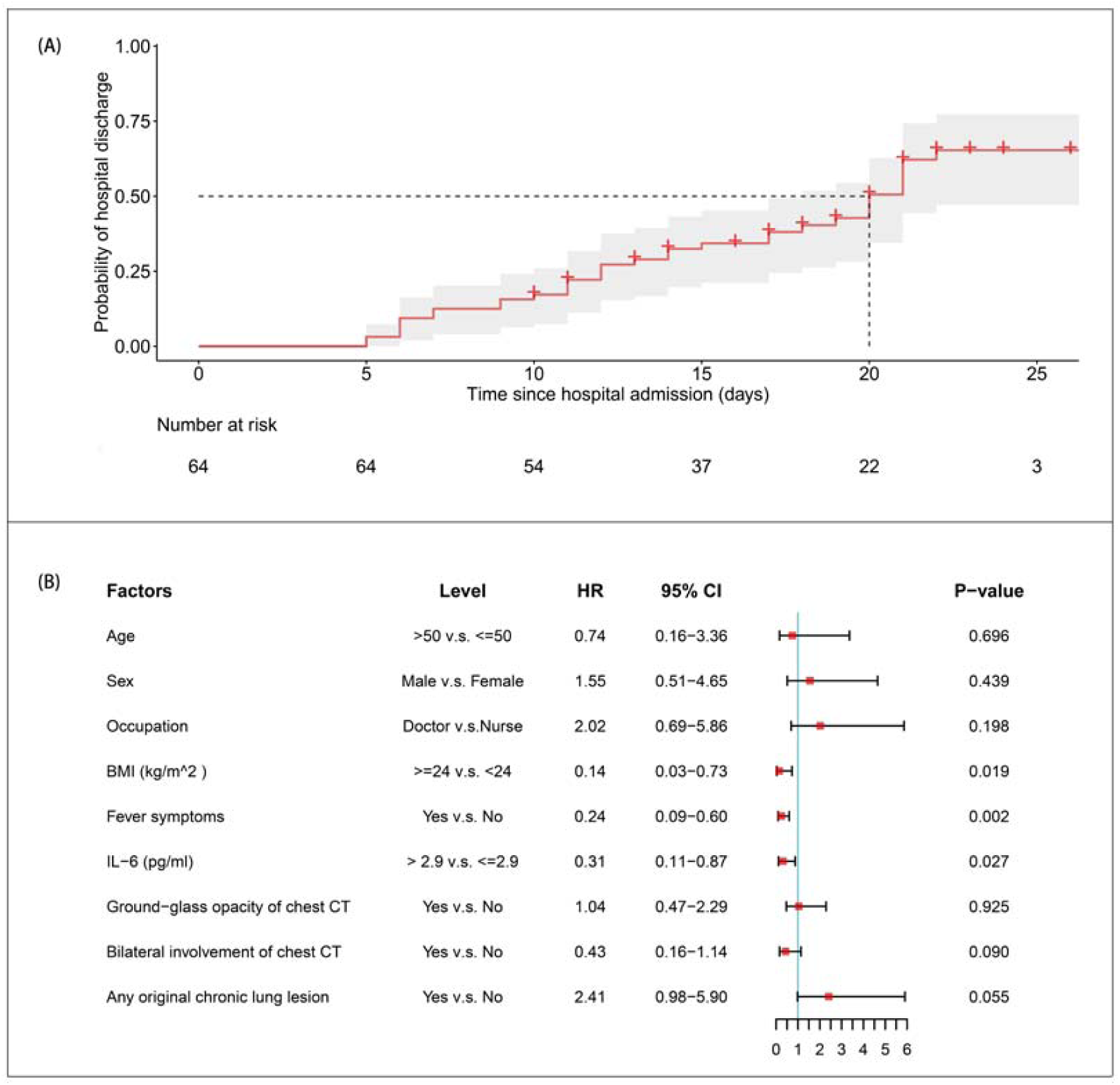
Hospital discharge rates and relevant risk factors of medical staff infected with COVID-19 pneumonia. (A) The probability of hospital discharge and the length of hospitalization. Gray area represents 95% CI ranges and symbol ‘+’ represents censored data. (B) The results of proportional hazard Cox model. Shown are estimated hazard ratios (HRs) and 95% CI ranges of age, sex, occupation, body mass index (BMI), fever, IL-6 levels, ground-glass opacity on chest CT, bilateral involvement of chest CT and any original chronic lung lesion. The endpoint of Cox model was hospital discharge and patients who remained in hospital as of 24 Feb 2020 were regarded as censored data.

It should be noted that the endpoint of Cox model was discharge, and patients who continued to be hospitalized as of 24 Feb 2020 would be regarded as censored data. The HR metric derived from multivariate Cox regression model was utilized to ascertain factors significantly associated with the endpoint of patients infected with SARS-CoV-2. Results of the final multivariate Cox regression model showed that larger BMI (≥ 24 kg/m^2^) (HR 0.14; 95% CI 0.03-0.73), fever symptoms (HR 0.24; 95% CI 0.09-0.60) and increased IL-6 levels (> 2.9 pg/ml) on admission (HR 0.31; 95% CI 0.11-0.87) were unfavorable factors for hospital discharge (all HRs <1 and all P-values <0.05) according to Cox regression mode (Figure 2B).

## Discussion

By 20 Feb, 2020, 476 hospitals across China had reported, in total, 2,055 laboratory-confirmed cases of medical staff with SARS-CoV-2 infection, of which the majority (88%) were from Hubei province [25]. According to China-WHO joint investigation report, most of the infections among medical staff occurred in the early stages of the COVID-19 outbreak in Wuhan, when there was a lack of knowledge about transmission approaches and experience to fight with the SARS-CoV-2 [25]. Despite the outbreak of COVID-19 occurring in few scattered hospitals (e.g. 15 medical staff were infected at one hospital in Wuhan), hospital-related transmission is not the main transmission feature of COVID-19 in China [25]. Our findings advocate this viewpoint. According to our data, a small proportion of 64 novel coronavirus-infected medical staff had a direct contact with specimens of patients (5%) as well as patients in fever clinics (8%) and isolation wards (5%) during patient care. In addition, none of the 64 medical staff had an exposure to Huanan seafood market or wildlife, and 4 (6%) had family members with confirmed COVID-19. The exact mode of medical staff infection remains unclear. The findings are consistent with previous reports [12, 25].

The demographic characteristics and clinical manifestations of medical staff with confirmed COVID-19 in Wuhan were not exactly the same as general confirmed patients included in recent studies [10, 12, 26]. In our study, most of the novel coronavirus-infected medical staff analyzed were females and nurses, and had a smaller median and range of age. The medical staff infected with SARS-CoV-2 have similar signs and symptoms with general confirmed infection patients [12, 26]. The infected medical staff tended to manifest on chest CT with bilateral, subpleural ground-glass opacities, which is consistent with the recent radiological reports on COVID-19 pneumonia [27-30]. Furthermore, abnormal D-dimer levels as well as abnormal functions of kidney, heart and liver was relatively rare among medical staff with SARS-CoV-2 infection.

In our study, only one of 64 medical staff with SARS-CoV-2 infection was severe case, none developed acute respiratory distress syndrome or transferred to intensive care unit. More than half of the cases were discharged by 24 Feb, 2020. Previous studies suggested that 13.8% of the general confirmed patients were severe cases, among whom older age, male sex, chronic diseases are more common [25, 31, 32, 33]. Contrarily, our study revealed that medical staff have relatively milder symptoms, which may be partly due to their medical expertise, younger age and less underlying diseases.

Medical staff with symptoms onset for less than 10 days by the time of admission were compared with those with symptoms of more than 10 days. We found that medical staff with symptoms for less than 10 days had more prominent laboratory abnormalities on admission, and they also experienced relatively worse clinical course and longer hospital stay. Furthermore, the median time between symptoms onset and admission of infected medical staff in this study was 8.5 days, longer than general population as described in recent publications [10, 21]. We believe that mild cases of infected medical staff without an early hospitalization was mainly because they made admirable concessions to provide the limited-number of isolation wards for infected patients with worse conditions during the peak time of COVID-19 epidemic in Wuhan.

Predictors of hospital discharge among infected medical staff were identified by Cox model. Smaller BMI, absence of fever and normal levels of IL-6 on initial stage were favorable factors for recovery and discharge. A recent study revealed that fever was identified in only half of the patients on presentation but increased to nearly 90% after hospitalization [26]. Elevated IL-6 levels were observed in 80% of infected medical staff on admission, which is associated with inflammatory response [34, 35]. To explore how absence of fever and IL-6 levels on initial stage affect the length of hospital stay and discharge of medical staff with SARS-CoV-2 infection, further studies are needed.

Given that epidemiology and clinical features of medical staff infected with SARS-CoV-2 is unclear, our study provides an insight to prevention and treatment of medical staff at risk of COVID-19 infection. So far, more than 40,000 medical personnel outside Hubei province gathered in Wuhan for the battle against the epidemic, and China has attached great importance to infection prevention among medical staff [25]. Although some safeguards have been introduced in the aspects including salary, injury suffered on the job, rest, and psychological adjustment for medical staff, the next step will continue to strengthen the promotion of these measures. Meanwhile, some potential problems remain to be solved, such as unclear patterns of infection, mental health care for medical staff [36], and the possibility of airborne transmission from aerosol production by medical practices in health care facilities [25]. A recent study from Singapore found that surface environmental and personal protective equipment contamination caused by respiratory droplets and fecal shedding from patients infected with SARS-CoV-2, suggesting that the environment is a potential viral vector [37]. Further investigations should be devoted to identifying the exact patterns of SARS-CoV-2 infection among medical staff.

## Limitations of this study

We acknowledge some limitations of this study. First, only 64 for medical staff with confirmed COVID-19 from a single hospital in Wuhan were included. However, the population from which they were sampled was large and we did not include all of the cases during the study period. In fact, there are 2,055 laboratory-confirmed cases of COVID-19 in medical staff as of 20 Feb 2020 [25]. This limitation in our study may result in deviations in epidemiological and clinical observation characteristics. We hope that the findings presented here will encourage a more comprehensive assessment of SARS-Cov-2 infection in for medical staff. Second, more detailed information, particularly regarding specific causes of SARS-CoV-2 infection among for medical staff, was unavailable at the time of analysis; however, this is a retrospective, observational study and the data used in this study only provide a preliminary insight into epidemiological features and clinical outcomes of a group of for medical staff confirmed with COVID-19. Further research on this regard is needed.

## Conclusion

The study medical staff have relatively milder symptoms and favorable clinical course, which may be partly due to their medical expertise, younger age and less underlying diseases. Smaller BMI, absence of fever symptoms and normal IL-6 levels on admission are favorable for recovery and hospital discharge for medical staff infected with COVID-19. Further investigations should be devoted to identifying the exact mode of COVID-19 among medical staff.

## Data Availability

Data are available from the corresponding author on reasonable request.

## Contributors

J.L., L.OY. and C.S.Z. conceptualized the article. J.L., L.OY., P.F., D.Y., Y.K.C., J.T. and S.Y.P. collected the epidemiological, clinical and radiological data. J.L., L.OY., P.G., H.S.W. and Y.L.C. cleaned and analyzed the data. J.L. and F.Y. contributed to radiological data interpretation. J.L., L.OY., P.G., H.S.W. and P.F. drafted the manuscript. X.Y.H., O.A., H.S.S., F.Y. and C.S.Z. revised the final manuscript. All authors approved the final draft of the manuscript. C.S.Z. is the guarantor. The corresponding author attests that all listed authors meet authorship.

## Declaration of interests

All authors declare no competing interests.

## Acknowledgments

We would like to thank all colleagues for helping us during the current study. The authors would like to express their appreciation for all of the emergency services, nurses, doctors, and other hospital staff for their efforts to combat the COVID-19 outbreak.

## Role of the funding source

This study was supported by National Natural Science Foundation of China (grant numbers: 81873919). The funder of the study had no role in study design, data collection, data analysis, data interpretation, or writing of the report. The corresponding authors had full access to all the data in the study and had final responsibility for the decision to submit for publication.

## Notes

### Competing Interest Statement

The authors have declared no competing interest.

### Clinical Trial

This is not a clinical trial.

